# Over 100 years of Rift Valley Fever: a patchwork of data on pathogen spread and spillover

**DOI:** 10.1101/2021.02.18.21251916

**Authors:** Gebbiena M. Bron, Kathryn Strimbu, Hélène Cecilia, Anita Lerch, Sean Moore, Quan Tran, T. Alex Perkins, Quirine A. ten Bosch

## Abstract

During the past 100 years, Rift Valley fever virus (RVFV), a mosquito-borne virus, has caused potentially lethal disease in livestock, and has been associated with significant economic losses and trade bans. Spillover to humans occurs and can be fatal. Here, we combined data on RVF disease in humans (22 countries) and animals (37 countries) from 1931 to 2020 with seroprevalence studies from 1950 to 2020 (N=226) from publicly available databases and publications to draw a more complete picture of past and current RVFV epidemiology. RVFV has spread from its original focus in Kenya throughout Africa and into the Arabian Peninsula. Throughout the study period, seroprevalence increased in both humans and animals, suggesting potentially increased RVFV exposure. In 24 countries animals or humans tested positive for RVFV antibodies even though outbreaks had never been reported there, suggesting RVFV transmission may well go unnoticed. Among ruminants, sheep were most likely to be exposed during RVF outbreaks, but not during periods of cryptic spread. We discuss critical data gaps and highlight the need for detailed study descriptions, and long-term studies using a one health approach to further convert the patchwork of data to the tale of RFV epidemiology.

## 1. Introduction

Over 100 years ago, in June 1912, an outbreak of “an obscure disease [that] caused heavy mortality in lamb” was described that, temporarily, had a discouraging effect on the sheep industry in Kenya [1]. Eighteen years later, in 1930, the likely causative agent, Rift Valley fever virus (RVFV), was isolated by Daubney and colleagues [2]. Nearly 100 years later, this primarily mosquito-transmitted virus, now known as Rift Valley fever phlebovirus [3], still causes morbidity and mortality in animals with spillover to people throughout the African continent. Outbreaks can lead to severe recurring economic losses, disrupting the livelihoods of often poor communities. Due to its economic impact, pathogenicity, and unpredictable (re)emergence, RVFV is recognized as a danger for both human and animal populations [4]. A better understanding of the eco-epidemiology of RVF could help inform intervention and surveillance strategies to reduce the burden of disease and minimize pathogen range expansion.

In the face of climate change, range expansions and redistributions of mosquito-borne viruses are expected [5]. RVFV, mostly transmitted by *Culex* and *Aedes* spp. mosquitoes, first expanded its range outside Africa in 2000, when it caused major outbreaks on the Arabian Peninsula [6–9]. The suitable geographical ranges for competent mosquito, and ruminant populations that are conducive to pathogen introduction and spread are changing, alarming Europe and the Americas [10,11]. Historically, the emergence of RVFV in new areas has been unpredictable, but it has frequently been linked to animal trade [12–14]. As “a transmissible disease that has the potential for very serious and rapid spread, without regards for national borders, and with serious socio-economic and public health consequence as well as major importance in the international trade of animals and animal products,” the World Organization for Animal Health (OIE) recognizes RVF as a notifiable animal disease of concern. Similarly, the World Health Organization’s research and development blueprint named RVFV a priority pathogen due to its “epidemic potential” [4].

The impact and disease burden of RVF on local economies and livelihoods exemplifies the potential devastating consequences RVFV could have on the global food supply and population health. Outbreak sizes in humans have been estimated to range from a few cases to thousands of cases, with case fatality risk ranging from 1 to 30% [15]. Human cases—ranging from mild to flu-like symptoms to hemorrhagic fever and death—are most common in individuals with close contact with animals and those consuming raw meat [16], often in rural communities. In Africa, more than 740 million individuals are currently living in rural, mostly agricultural communities, and this is expected to increase to 1,039 million by 2050 [17], placing them and their animals potentially at risk for RVF. Outbreaks in animals are recognized by abortion storms and high mortality in young animals [2]. Although sheep and goats are most susceptible, RVFV also affects cattle, camels, and, sporadically, wildlife [18,19]. No RVFV treatments are registered for humans or animals, and medical treatments, when available, are limited to supportive care.

The strategies for RVF outbreak prevention and control are limited [20]. At a local level, communities and livestock owners could implement strategies to reduce mosquito bites, e.g., mosquito-control, moving animals to high-elevations pastures during peak mosquito seasons. In addition, individual animals can be protected and herd immunity can be accomplished by vaccinating animals [21,22]. However, preventative vaccination can be challenging due to a low burden of disease in the absence of outbreaks, logistical and economical barriers, in part due to limitations of the currently available vaccines. For example, the most commonly used vaccine in livestock is a live-attenuated vaccine (i.e., Smithburn vaccine) which is highly effective [23], but can cause abortion and birth defects when vaccinating pregnant animals [24]. When RVFV is suspected in animals, control measures include announcements for hygiene measures and awareness [25]. However, when cases are confirmed in humans, a widespread outbreak is generally suspected and more expensive and intense measures are used, ranging from vector control to trade bans [25,26]. These interventions can disrupt local, regional, and national economies. For example, the estimated economic impact of a RVF outbreak in 2006-2007 ranged from 0.01% of the gross domestic product (GDP) in Tanzania (6.7 million US$) to 5.5% of the GDP in Somalia (471 million US$), making outbreak prevention a potentially more cost-effective strategy [27,28]. To strengthen RVF outbreak prevention, predictive modeling could be used to facilitate early community communication, targeted mosquito control interventions, and even localized vaccination campaigns [29].

RVF outbreaks in animals and humans are associated with a variety of factors, but the eco-epidemiology is not fully understood. RVFV circulates through different transmission routes and infection may occur through an infectious mosquito bite, or by contact with infected tissues and fluids (e.g., aborted tissues, exposure during slaughter). Outbreaks in animals, and subsequently in people, are often associated with increased rainfall in historically endemic areas, most notably the foci along the Great Rift Valley [30,31]. The virus, capable of vertical transmission, likely survives in *Aedes* spp. eggs in the environment. Rainfall and flooding subsequently allow for virus re-emergence. Next, flooded areas provide ample breeding habitat for mosquitoes, further amplifying transmission. These patterns have been seen during, for example, the 1973-75 South African, and the 1997-98 and 2006-2007 Eastern Africa outbreaks, which were associated with El Niño rain events [32,33]. In other areas, RVFV may be introduced via the live-animal trade. Movements of infected animals from endemic regions with year-long presence of mosquitoes, can (re)introduce the virus to more seasonal ecosystems, in a source-sink fashion. Trade-related introductions and outbreaks are frequently associated with large gatherings, whereby large numbers of, potentially infected, animals are brought in for slaughter [12,34,35]. Despite the recognition of the underlying factors, the complexity of the dynamics together with a scarcity in data hamper actionable risk assessments.

To improve our understanding of the historic, current, and future epidemiology of RVF, and to aid in planning future surveillance and intervention strategies, we compiled historical RVF case data and seroprevalence studies of both humans and animals. We stitched a patchwork of the wide range of available RVF data sources (including grey and white literature) together to examine patterns associated with outbreaks, the relative contributions of ruminant species to RVFV outbreaks and cryptic spread, and ultimately spillover to humans.

## 2. Materials and Methods

To explore temporal and spatial activity of Rift Valley fever virus (RVFV) in Africa and beyond, we combined human and animal (domesticated animals and wildlife) case records and serological studies. To create a data set as complete as possible, we included information sources beyond peer-reviewed (experimental) studies (an integrative literature review), but refrained from expert consultation and interviews. We extracted data from case reports, outbreak reports, and serological studies available in openly accessible, online databases, and in peer-reviewed journals. We then explored relationships among these records. Mosquito records were excluded.

### 2.1. Data acquisition strategy

Data was extracted in five steps, A to E, by hand (Figure 1). First we obtained RVF case data from databases on outbreaks in humans and animals (Step A), complemented with data from systematic literature reviews (Step B). Next we extracted seroprevalence data from systematic literature reviews by cross-referencing included studies (described in more detail later) (Step C). We updated existing reviews to the current date (Step D) and confirmed the absence of RVFV evidence for selected countries (Step E, Figure 1). The database was last updated on January 13th, 2021, and includes publications published up until December 31st 2020, including data pertaining to 2020 and earlier.

**Figure 1.**
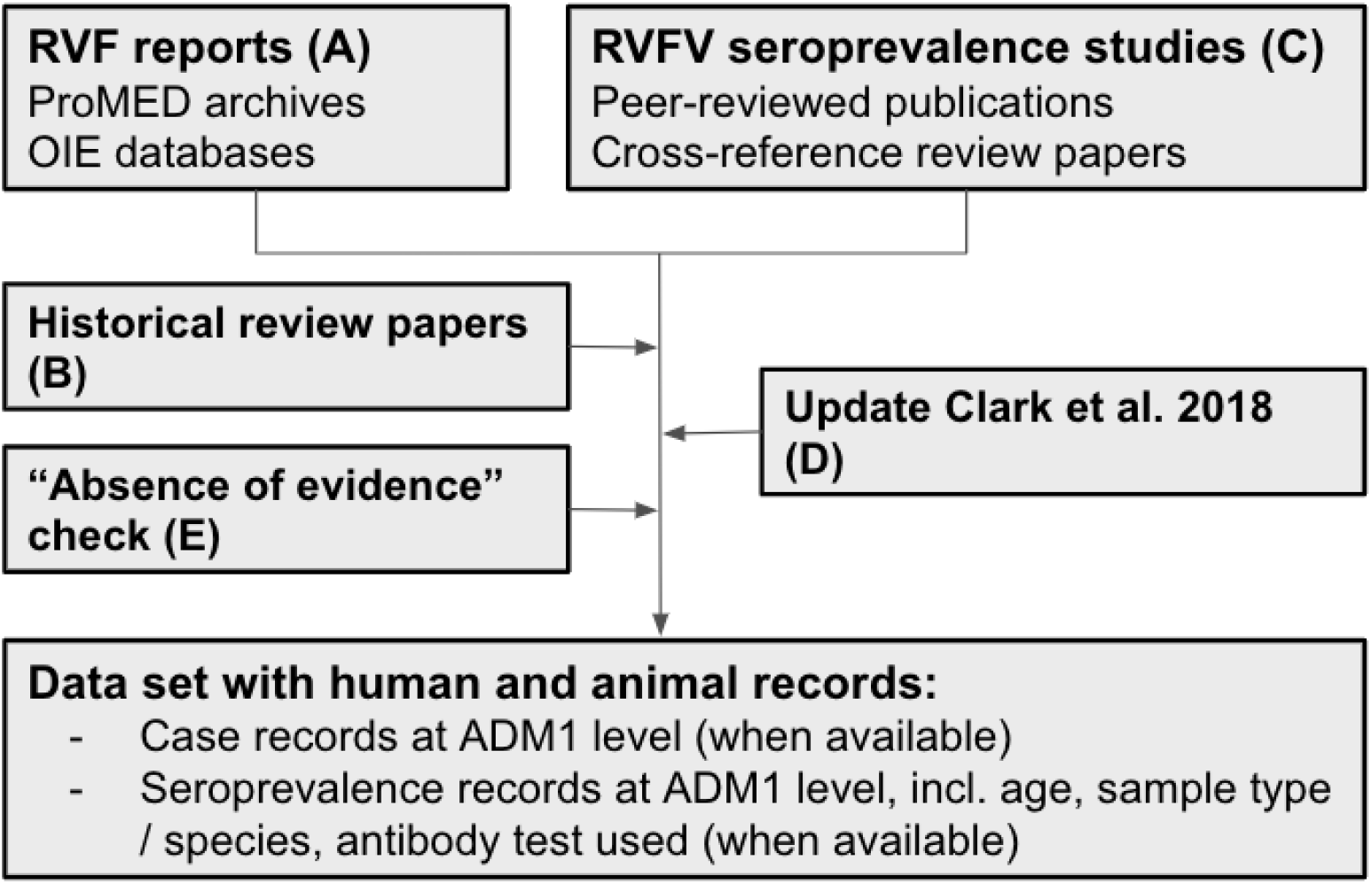
Flowchart of the source and data acquisition. A total of 275 sources (A: two OIE databases and ProMED, and B to E: 272 publications) were used to inform the RVF case and RVFV seroprevalence data set for humans and animals. RVF reports were extracted from databases (A) and supplemented with historical review papers (B). Seroprevalence data was first extracted from publications identified through cross-referencing review papers (C) and the search strategy of Clark et al. 2018 [45] was repeated to identify and add publications from 2017 to 2020 (D). In step E, a search was conducted to explore if publications were available for African, Southern and Western Asian countries without RVF reports and RVFV seroprevalence studies to confirm absence of RVFV evidence. Data was included until December 31^st^ 2020, the last update was conducted January 13^th^ 2021 by repeating step A and D.

In step A, RVF case records were extracted from posts from the Program for Monitoring Emerging Diseases (ProMED) from the International Society for Infectious Diseases (ISID) and the public databases from the World Organization for Animal Health’s (OIE). ProMED was established in 1994 and archived posts were accessible from 1996 onward [36]. The OIE’s Handistatus 2.0 database [37] contained data from 1995 to 2004, after which it was replaced by the current World Animal Health Information System (WAHIS / WAHID) interface [38]. RVF case data were compared against the World Health Organization’s Rift Valley fever outbreak bulletins [39] and the Centers for Disease Control and Prevention (CDC) outbreak summaries [40] from 2000 onward to ensure no human RVF cases were omitted. Next, in step B, additional case records were extracted from historical review papers and reports [33,41–44].

To identify and extract data from original seroprevalence studies we built on existing systematic literature reviews [45,46], and cross-referenced references within these publications (Step C). Hereby, we included all geographical regions, e.g., studies from outside Africa and the Arabian Peninsula were included. Next, we used the search strategy from the most recent RVFV seroprevalence review, Clark et al. (2018) [45], to include publications published after 2016. Using the search term ((“Rift Valley Fever” OR “rvf”) AND (“prevalence OR “incidence” OR “sero)) in PubMED and Web of Knowledge, 102 and 162 publications were identified respectively. There were 88 duplicates between the two searches. Of the 176 unique publications, 34 were already in our data set (i.e., obtained through cross referencing), 88 were excluded based on their title (e.g., not relating to RVF), for 54 publications the abstract was assessed, 21 were excluded as they did not contain information pertaining to RVF or did not include original or new data, and data from 33 publications were added to the data set. In addition, we conducted an absence of evidence search (step E) to explore if publications were available for countries form African, Southern and Western Asian United Nations (UN) regions [47] with neither serological nor outbreak data (Google Scholar search terms: (“*country name*”, “Rift Valley fever virus”, serology)). For 30 countries no RVFV data was found, including six countries on mainland Africa: Algeria, Burundi, Eritrea, Guinea Bissau, Lesotho and Liberia.

### 2.2. Data set

Information parsed from the different sources was summarized in four data types: human RVF cases, animal RVF cases, human serology, and animal serology.

#### 2.2.1. RVF case definition

Case definitions for RVF vary between agencies, reporting countries, and reports. As such, we did not use a standardized case definition and included any reported and suspected cases. Broadly, a case is defined as an individual infected with RVFV, with or without clinical symptoms. Cases of RVF in animals are often recognized by clinical symptoms in the herd, but may go unnoticed. Determining the number of infected animals is therefore challenging. The OIE includes records on suspected transmission without case confirmation in their HandiStatus2.0 and WAHIS database. We included those entries in our animal RVF data set as well. In contrast, a human case of RVF is often defined as an individual with moderate to severe clinical symptoms who sought medical care. For both animals and humans, case numbers were included in the data sets, when available, but again it is important to note that these cases represent varying levels of certainty. Cases were not always associated with a confirmatory diagnostic test.

A year with RVF cases was defined as any calendar year for which positive case numbers in humans or animals were reported or suspected for a country. We use this term synonymously with RVF outbreak, as an outbreak year was defined as any calendar year for which case data were reported. Laboratory-based cases are not considered. Inter-epizootic cases were included, whereas travel-related cases were excluded. Sometimes, an outbreak year only had one reported case. RVF outbreaks that spanned two calendar years (e.g., November to February) were marked in both years. When data sources reported a year from July to June (sometimes referred to as collating by season) and no additional information was available about the time of the observation (i.e., month or week), we also marked the observations in both calendar years.

#### 2.2.2. Geographical information

Countries were grouped according to UN geographical regions [47]. The Canary Islands (ES-IC) were grouped with Northern Africa. Countries were abbreviated using three-letter codes and, when possible, information was parsed to administrative level 1 (ADM1) using the regions, districts, and provinces included in the Database of Global Administrative Areas (GADM) [48]. ADM2 and other smaller locations referenced (e.g., farm names) were collated at the ADM1 level. Since our database spans 91 years, including the period of decolonisation in Africa, when many nations (re)gained their independence, country names changed, borders were adjusted, and new nations were formed. To address this, we matched countries with the current UN and GADM database (e.g., Rhodesia, now Zimbabwe). In addition, historic RVF cases and serological studies may have taken place in an area of a country which is now recognized as an independent country by the UN. We used the current country names based on ADM1 level information or general regional directions of the data (e.g., South Sudan). Hence, older studies can be linked to countries that were not formally established at the time the study was conducted.

#### 2.2.3. Serological records

Reports on the presence of RVFV antibodies in humans and animals were summarized by country (ADM0), ADM1 (when available), year(s) of sample collection (if this information was not available authors were contacted), number of individuals tested, number positive, and study sample characteristics. Data were further characterized including age of human participants, animal species, or people sampling strategies (e.g., random sampling of a general population; sampling of febrile, hospitalized or suspected patients; testing of high-risk individuals; unclear strategies or mixed samples), the type of antibody test used (e.g., enzyme-linked immunosorbent assay, immunofluorescent antibody test, virus neutralization test, complement fixing, plaque reduction neutralization tests), and antibody type targeted. If both IgM and IgG results were available, we included the IgG results and marked IgM availability in the notes. Note: the presence of IgG antibodies does not mean virus is circulating in the area where the sample was taken, movements and vaccination status of individuals should be accounted for. Cross-reactivity with other circulating viruses may also interfere with the interpretation of seroprevalence estimates, depending on the test used. In areas without known cases the presence of RVFV should be confirmed, and paired sera should, ideally, be taken.

### 2.3. Statistical analyses

Summary statistics regarding the temporal and spatial extent of each of the data sets are presented first. To further explore the aggregated data at the country and year level, we assessed how RVFV seroprevalence, and case data of animals and humans were associated to each other in regression analyses.

#### 2.3.1. Data preparation

For our statistical analyses we aggregated data at the country level, because for about a quarter of the entries ADM1 was unknown (397 of 1511 records). Seroprevalence data were cleaned prior to analysis, excluding IgM-only records, records with missing data (e.g., study year or exact number of positives), and data points that could not be categorized as either outbreak or non-outbreak associated (e.g., data points reporting multiple years only partly overlapping an outbreak). A serological record was considered associated with RVF outbreaks if the samples were taken during the year in which RVF cases were recorded in humans or animals in the same country, or in the year after. Here, we made the assumption that post-outbreak serology is typically performed in the outbreak region. We test this assumption on the subset of data for which sufficient information is available. For those instances that this cannot be verified, we discuss the impact of the assumption. We limited outbreak-associated serology to those surveys performed up to one year post outbreak.

#### 2.3.2. Annual RVF case and serological study availability

We estimated the increase in annual data availability for the four data types to quantify the progress made in data availability over time. For each data type, the response variable denoting if RVFV cases or a serological study was available in a given year (1) or not (0) was regressed against year: 1930 to 2020 for RVF cases and 1930 to 2017 for serological studies.

#### 2.3.3. Variation in seroprevalence

We compared seroprevalence between samples that were or were not associated with recent RVF cases (outbreak-associated: yes or no) using logistic regression models weighted by sample size. In these models, we accounted for differences between geographic regions (reference level: Eastern Africa) and over time. Time was rescaled to ten-year time steps and the start year of sample collection was used when a study reported multiple sampling years combined. The regression analysis of human seroprevalence data was conducted for a subset of data including randomly sampled individuals only. The regression analysis of animal seroprevalence data was conducted for the full data set and a subset of data including ruminant samples only (including camels, but excluding mixed samples with horses, mixed wildlife samples, and donkey, pig and rodent samples). To further dissect possible species contribution to RVFV, the logistic regression was repeated on a subset of data including only records for single ruminant species (sheep - reference level-, buffalo, camels, cattle, goats). The model accounted for difference in time and geographical region, as above, and assessed the association of species, recent RVF cases, and their interaction, with seroprevalence. Variance inflation factors were assessed (<3 was accepted). Model estimates were converted to odds ratios representing the adjusted odds ratios (aOR) of finding a positive sample in the group compared to the reference group.

Lastly, we extracted all seroprevalence data when RVF had never been reported in the country. We compared, by ANOVA, if mean seroprevalence (by country and year) was different based on the current, 2020, RVF status of the country (i.e., RVF cases have been reported since the seroprevalence study was conducted, or no cases have been reported to date).

#### 2.3.4. Concurrent animal and human RVFV activity

To assess if animal case investigations were more likely when human RVF outbreaks occurred, we compared, by Fisher’s exact test, if the proportion of animal case records with case counts was different when human outbreaks had or had not occurred in the same year and country. To determine if countries with higher seroprevalence in animals were more likely to have higher human seroprevalence, we summarized seroprevalence by country and year for randomly collected human samples and ruminant animal samples, and assessed the Spearman’s rank-based statistic. As above, we acknowledge that the spatial scale of RVF outbreaks is typically smaller than the country level. We further examine the limited amount of studies for which ADM1-level information is available for both human and animal studies to answer two questions: i) how likely are studies in humans and animals from the same year to have been performed in the same ADM1, and ii) for those records, can we distinguish patterns between human and animal seroprevalence and are those consistent with conclusions from the larger dataset?

#### 2.3.5. Software

All statistical analyses were conducted in R Statistical Computing Software [49]. Package *lme4* [50] was used for regression models. Figures were created using packages *rnaturalearth* [51] and *ggplot2* [52], and organized using *cowplot* [53].

## 3. Results

Information from OIE databases and ProMED archives was supplemented with data from 272 peer reviewed publications (Table 1). The data set consists of 1,509 records of four data types: 125 on human RVF cases extracted from 59 sources, 415 on RVF cases in animals from 38 sources, 294 on human serology from 106 sources, and 675 records on animal serology extracted from 143 sources (Suppl. Table 1).

**Table 1.** Peer-reviewed publications by region. The median year of publication and range are included. The OIE databases (n=2) and ProMED archive (available since 1996) are not included in this table.

| Continent | Region | Sources | Year of publication of used sources |  |  |
| --- | --- | --- | --- | --- | --- |
|  |  |  | Earliest | Median | Most recent |
| Africa | Central | 22 | 1965 | 2008 | 2019 |
|  | Eastern | 102 | 1931 | 2013 | 2020 |
|  | Northern | 35 | 1978 | 1999 | 2020 |
|  | Southern | 33 | 1951 | 2011 | 2020 |
|  | Western | 46 | 1980 | 2010 | 2020 |
| Asia | Southern | 2 | 1995 | 2000 | 2005 |
|  | Western | 21 | 1984 | 2013 | 2020 |
| Multiple and other regions* |  | 11 | 1969 | 2011 | 2020 |
| **Total** |  | 272 | 1931 | 2011 | 2020 |

\* One publication from Poland, Europe, is available [54].

### 3.1. Data availability

#### 3.1.1. Data availability over time

Ninety-one years were included in the data set (1930 to 2020), with the first publication in 1931 [2] (Table 1, Figure 2). Publications on human and animal RVF cases were first available in 1931. The first publication on human seroprevalence estimates was published in 1956 [55], and on animal seroprevalence estimates in 1958 [56]. The average lag between study-end year and publication year of the manuscript for seroprevalence studies was 3 years for both human and animal studies. RVF cases that occurred prior to 1990 often had a lag in their publication, as case records were published in peer-reviewed sources (e.g., historical reviews, compared to current near real-time reporting). During the 1990s, this transitioned to predominantly same-year reporting. Over 40% of human and animal RVF reports we extracted came from archived ProMED posts (37 of 89 human, and 125 of 279 animal RVF records published after 1996, respectively), which were mostly reported the same year as the outbreaks occurred.

**Figure 2.**
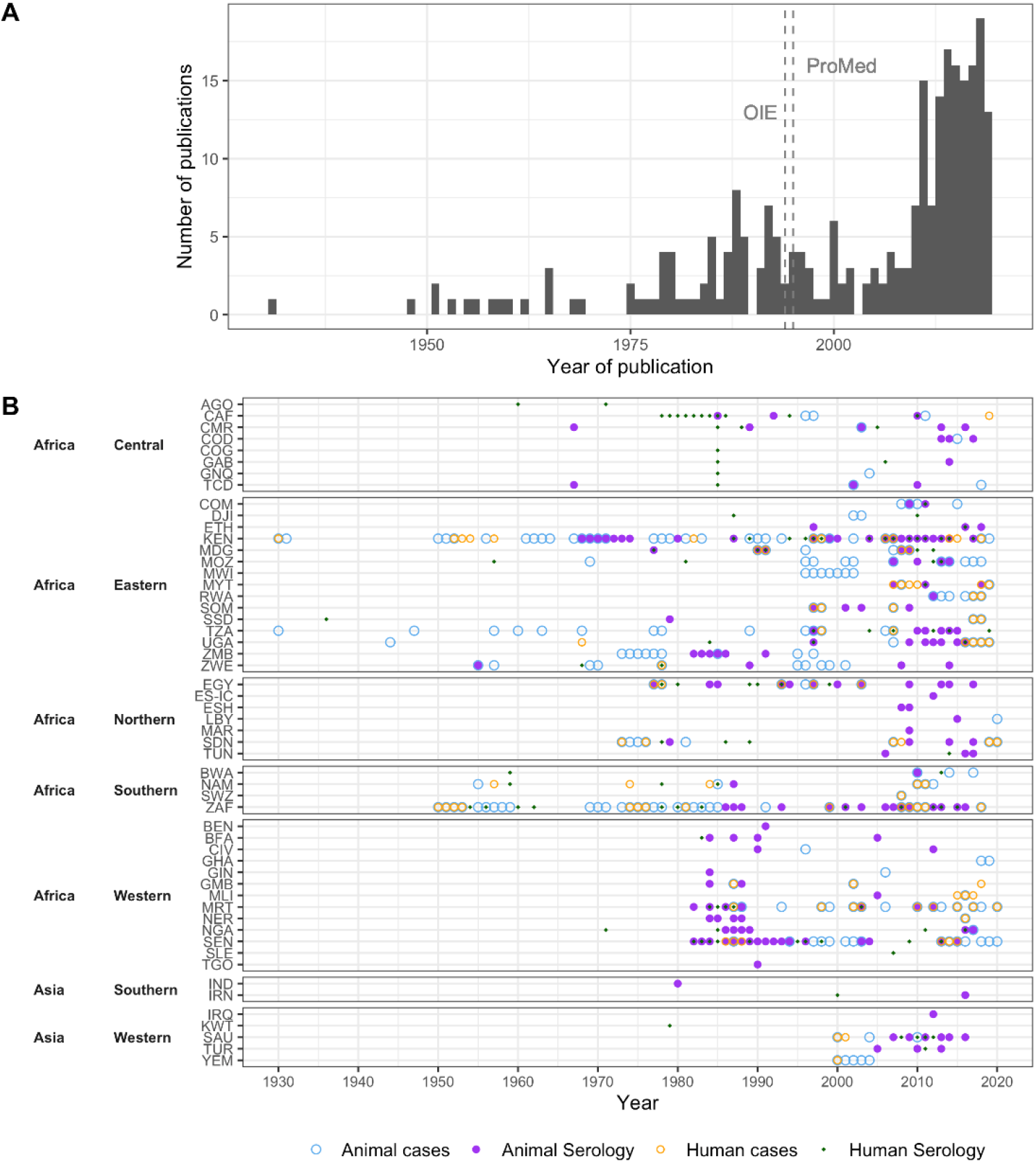
Regional variation in RVF reports and seroprevalence studies over time. **A)** Number of included publications by year. Vertical lines represent the first year during which OIE and ProMED reports were included. **B)** For each country, the years during which Rift Valley fever cases were reported, and the years during which Rift Valley fever virus seroprevalence data were collected are marked for animals and humans.When results from multiple years were reported as one, e.g. 1991-2000, we marked the first year in the figure. Serological data also include studies where no RVFV antibodies were detected, note that this is not a RVFV detection chart.

The number of RVFV publications increased over time (Figure 2A). Specifically, the probability of there being at least one record in a given year for each of the four data types increased significantly (P<.001), although the rate of increase differed between data types. This increase in publications was steepest for animal serology (beta = 0.124, 95% Confidence Interval [CI]: 0.082, 0.184) in comparison to human serology (beta = 0.089, 95%CI: 0.058, 0.129), human RVF cases (beta = 0.054, 95%CI: 0.033, 0.079), and RVF cases in animals (beta = 0.064, 95%CI: 0.038, 0.098) (Suppl. Figure 1).

#### 3.1.2. Spatial distribution of data

All five African regions, and about 80% of African countries, were represented in the data set and thus had either RVF cases reported or seroprevalence studies conducted (Figure 3). In addition, Southern and Western Asia were included with two (Iran and India) and five countries (Saudi Arabia, Yemen, Iraq, Kuwait, and Turkey), respectively (Figure 3). Of the 52 countries in the data set, human and animal RVF case records were found for 22 and 37 countries, respectively (Figure 3C and D). Human and animal seroprevalence studies were conducted in 33 and 45 countries, respectively (Figure 3E and F). Most human and animal serological publications (N= 106 and N=142) originated from Kenya (22 and 15) or South Africa (10 and 13), followed by Egypt (7 and 12). For 15 countries both human RVF cases and human seroprevalence data were available. For twice the number of countries (31 countries) both animal RVF cases and animal seroprevalence have been reported.

**Figure 3:**
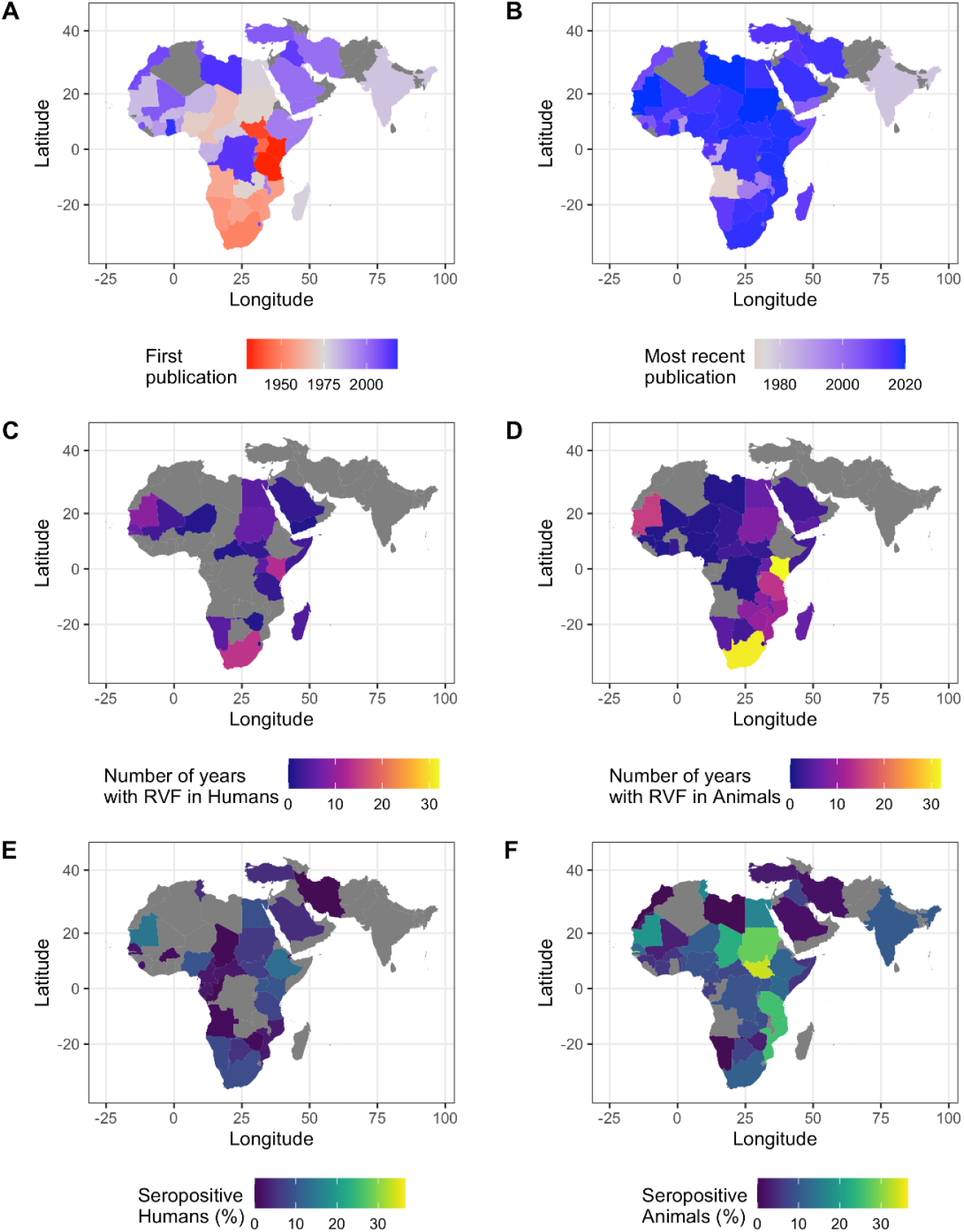
RVFV reporting and activity. **A and B)** The year of the first (A) and most recent (B) RVFV publication by country. The studies on the Canary Islands and Mayotte are not shown. No publications were found for the countries in grey. **C and D)** Number of years with human (C) or animal (D) RVF case records per country. **E and F)** Percent of individuals positive for RVFV antibodies of all individuals sampled per country for human samples (E) and animal samples (F). All samples, except IgM-only records were included. Gray indicates that no RVF case reports, or seroprevalence studies were found for this country. It should be noted that the presence of individuals with a positive RVFV antibody test does not ascertain local circulation of the virus and should be interpreted with caution, particularly for countries with no confirmed cases.

### 3.2. RVF outbreaks and number of cases affected

A total of 605,005 animal and 10,923 human RVF cases were reported during 68 and 45 years, respectively (note, rough estimates were not included in these totals). Animal and human cases were most often reported in Kenya (32 and 13 years, respectively) and South Africa (31 and 15 years, respectively) (Figure 3 C,D). Most animal cases were, also, reported from Kenya (507,996, 84% of total cases), followed by Tanzania (38,167) and South Africa (19,543). Most human cases were reported from Sudan (2,534), Egypt (1,267) and Kenya (1,187). A total of 224 animal and 96 human RVF case records were available after aggregating data by country and year. For 150 of the 320 aggregated records case counts were available (46.8%). Most of the records with case counts (126 of 150, 84.0%) occurred in 2000 or later. The number of RVF cases in animals ranged from a single animal succumbing to RVFV (e.g., an antelope in Senegal in 2020) up to estimates of 250,000 animals affected during the 1950–1951 outbreaks in Kenya. Human case counts also ranged from one individual to about 1,500, but it was noted that an estimated 20,000 to 100,000 people may have been infected in South Africa in the 1950s.

### 3.3. Variation in seroprevalence

Over 250,000 individuals (human and animals) were represented in the seroprevalence data and 210,074 of which met the inclusion criteria: 131,039 animals and 80,406 humans. The percentage of individuals that tested positive for RVFV antibodies by country and year ranged from 0% to 40% in humans, and 0% to 70% in animals (Figure 4A and C).

**Figure 4:**
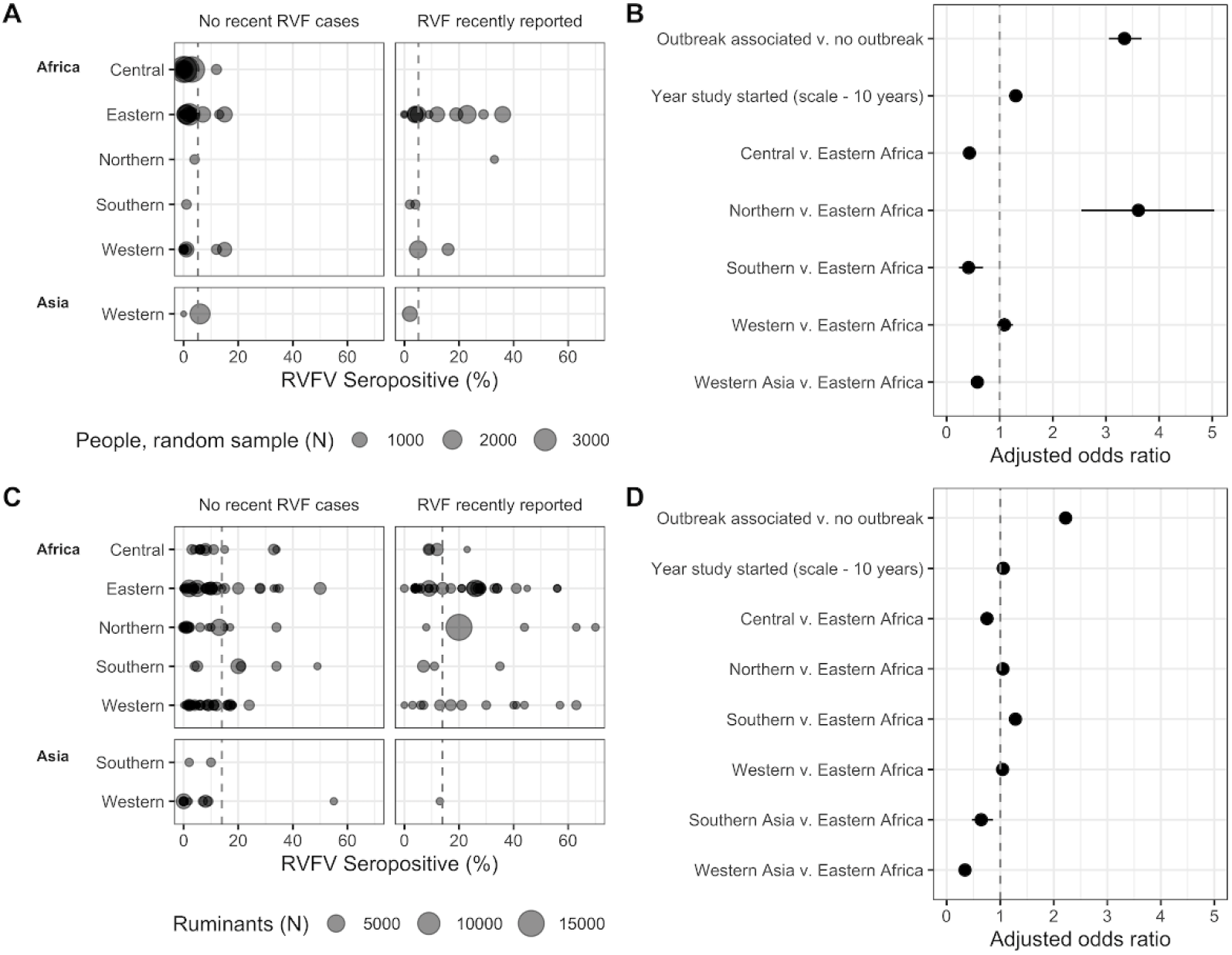
Regional human (A, B) and ruminant (C, D) RVFV seroprevalence in the absence of recent human and/or animal RVF cases or within the same year, or year post, reported RVF cases. **A and C)** Sample size and seroprevalence per country and year. The grey, dashed vertical line represents the overall mean seropositivity in humans (A) and ruminants (C). Note: dots can overlap. **B and D)** Adjusted odds ratio for odds of detecting a seropositive person (B) or ruminant (D). The grey, dashed vertical line represents equal odds. Horizontal lines represent 95% confidence intervals around the point estimate. Southern Asia was represented by studies from Iraq and India, Western Asia was represented by Iran, Saudi Arabia, Turkey and Yemen.

#### 3.3.1. The association of seroprevalence with time, region, and outbreaks

A subset with data on 48,702 randomly selected human individuals remained, after excluding those studies that targeted individuals with high-risk lifestyles and professions, febrile patients, and studies that had mixed or unclear sample selection. Similarly, 122,080 ruminant samples remained (including camels), after excluding studies with non-ruminants (e.g., rodents, horses, pigs, donkeys), and mixed samples of ruminants and non-ruminants (e.g., “pig and sheep”).

Among all included samples, 5.2% human samples and 14.0% of ruminant samples were positive for RVFV antibodies. Of samples associated with an outbreak (i.e., human and/or animal cases in the same or previous year) 12.6% of human samples and 19.9% of ruminant samples were positive for RVFV antibodies. This was substantially lower for samples not associated with outbreaks, with 2.7% of human samples and 9.8% of ruminant samples positive for RVFV antibodies (Figure 4A, C). Indeed, when assessing the subset of randomly selected individuals, the adjusted odds (aOR) of finding a positive serum sample was 3.35 times higher (95%CI: 3.06, 3.67, P<.001) when sampling was conducted in the same year or the year after RVF cases were reported in the country compared to sampling conducted in the absence of recently reported RVF cases (Figure 4B). A similar, yet smaller effect was observed for ruminants, with aOR=2.22 (95%CI: 2.14, 2.30, P<.001), indicating that more individuals get exposed to the virus during outbreak years than during cryptic cycles (Figure 4D). The proportion of individuals who tested positive for RVFV antibodies also varied over time and by region (Figure 4). Every ten years, the probability for a sample to be positive increased 1.30 times (95%CI: 1.24, 1.37, P<.001) for the subset of randomly selected humans and, similarly, 1.06 times per ten-year timestep for ruminants (95%CI: 1.04, 1.07, P<.001) (Figure 4B, D). In addition, relative to Eastern Africa, the proportion of individuals who tested positive was higher in people in northern Africa (aOR: 3.61, 95%CI: 2.53, 5.04) and lower in individuals from central and southern Africa and western Asia (i.e., Kuwait, Saudia Arabia, Turkey) (aOR: 0.43, 0.41 and 0.58, 95%CI: 0.36, 0.51; 0.23, 0.69 and 0.49, 0.68)(Figure 4b, Suppl. Table 2). The probability for a ruminant to be seropositive in middle Africa, and southern (i.e., Iran and India) and western Asia (i.e., Iraq, Saudi Arabia, Turkey) was lower than in eastern Africa (Figure 4D, Suppl. Table 3). The seropositivity in the ruminant populations in north, west, and south African regions was similar to slightly higher than, eastern Africa. Observed patterns of time and outbreaks on the proportion of individuals who tested positive in ruminants were robust to adding non-ruminants to the data set (Suppl. Table 4). For most records, it was not possible to verify if the seroprevalence studies were performed in the same ADM1-level as where the outbreaks occurred. About half of human outbreak records (65 of 125), and about a third of animal outbreak records (135 of 415 records) did not have ADM1-levels reported. When looking at the seroprevalence subsets for which ADM1-level were available (77.7% of randomly selected humans records [32,434 individuals], and 80% of ruminant record [78,773 individuals]), and taking a conservative approach to considering a sample outbreak associated (i.e., if an outbreak occurred in the country, and the ADM1 was not known, the sample was not outbreak associated) the general conclusions are robust, though effect sizes vary, and some regional associations changed (Suppl. Table 5 and 6).

**Table 2.** **Species specific seroprevalence** and the adjusted odds ratios for interaction between species and recent RVF reporting. Common livestock species were compared to sheep tested when RVF had and had not been recently reported in a country. The logistic regression model also accounted for the year the study was started and the region the work was conducted in (Suppl. Table 7). aOR: adjusted odds ratio; CI: confidence interval.

| Species | No recent RVF cases |  |  |  | RVF recently reported |  |  |  |
| --- | --- | --- | --- | --- | --- | --- | --- | --- |
|  | % | (N) | aOR | 95% CI | % | (N) | aOR | 95% CI |
| Sheep | 8.1 | (17,328) | 1 | (Reference) | 22.1 | (10,011) | 3.14 | (2.91, 3.39) |
| Buffalo | 13.8 | (1,417) | 1.49 | (1.26, 1.76) | 15.7 | (1,523) | 1.89 | (1.19, 2.99) |
| Camels | 10.0 | (4,938) | 1.81 | (1.61, 2.03) | 20.5 | (1,451) | 2.06 | (1.84, 3.90) |
| Cattle | 13.0 | (22,288) | 1.48 | (1.38, 1.59) | 18.2 | (19,519) | 2.16 | (1.70, 2.74) |
| Goats | 11.9 | (8,746) | 1.25 | (1.14, 1.36) | 17.1 | (5,968) | 1.85 | (1.39, 2.46) |

#### 3.3.2. Variation in exposure of ruminant species

The percentage of animals testing positive varied between the different ruminant species sampled, with sheep having the lowest seroprevalence in the absence of recent RVF cases (Table 2). However, the interaction between species and outbreak-associated sampling was strongly significant (P<.001). While the odds of detecting a seropositive sheep in association with recent RVF cases more than tripled, the odds of finding positive buffalo, camels, cattle and goats only increased 1.26 to 1.48 times (Table 2, Suppl. Table 7). The higher odds of detecting a positive sheep in association with RVF cases, indicate that this species could be more likely to be exposed during outbreaks, possibly amplifying the outbreak.

#### 3.3.2. RVFV activity prior to RVF case reporting

Serological studies took place in 27 countries (including the Canary Islands) without known RVF cases, for a total of 50 aggregated records by country and year (Figure 5). In 24 countries animals tested positive for RVFV antibodies; in 11 of these countries RVF cases have never been reported to date, whereas in 13 countries RVF cases were reported 1 to 38 years later. Notably, animals tested positive for RVFV antibodies in Iraq, Iran, and Turkey (Figure 3). However, confirmed animal or human cases have not been documented in these countries to date.

**Figure 5:**
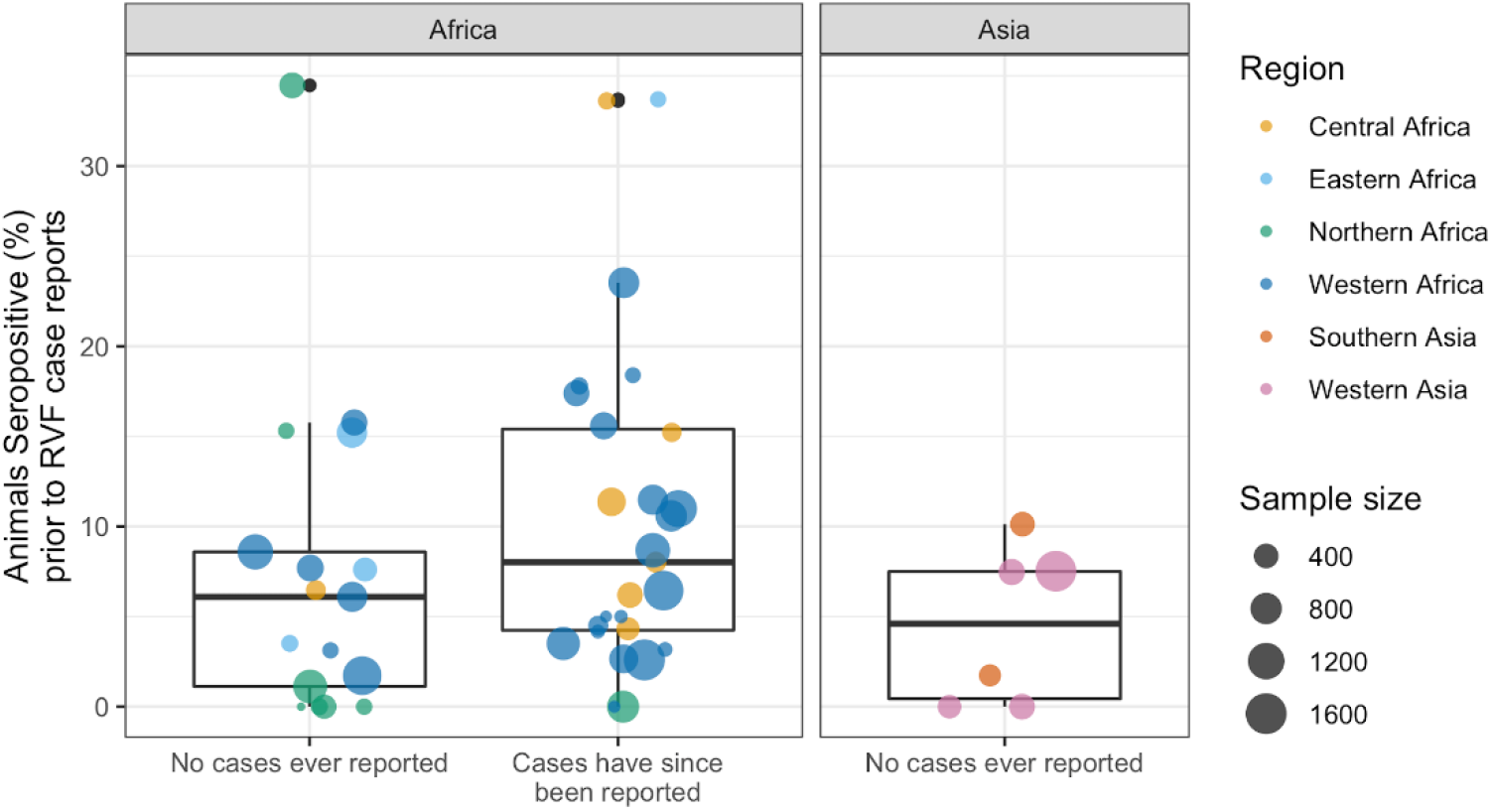
Rift Valley fever virus seroprevalence when RVF cases had never been reported in the country (human and/or animal). The outliers represent samples from Tunisia, no known RVF cases, during 2017-2018 (34.5% positive of 470 camels) [57], and two countries currently known to report RVF cases, South Sudan in 1979-1983 (33.7% positive of 92 ruminants, cattle and goats) [58], and Cameroon in 1968 (33.6% of 122 sheep) [59].

A total of 1,859 of 24,652 animals tested positive for RVFV antibodies in countries without known RVF cases at the time of the survey (7.5%), 8.8% of animals from countries that later reported RVF cases (1,217 of 13,788) and 7.2% of animals sampled in countries that, to date, have not reported RVF cases (827 of 11,454, Figure 6). No difference in seroprevalence, by country and year, was detected between the groups that currently report and do not report RVF cases (ANOVA, df 1,48, F=2.5, P=.12, Figure 5).

**Figure 6:**
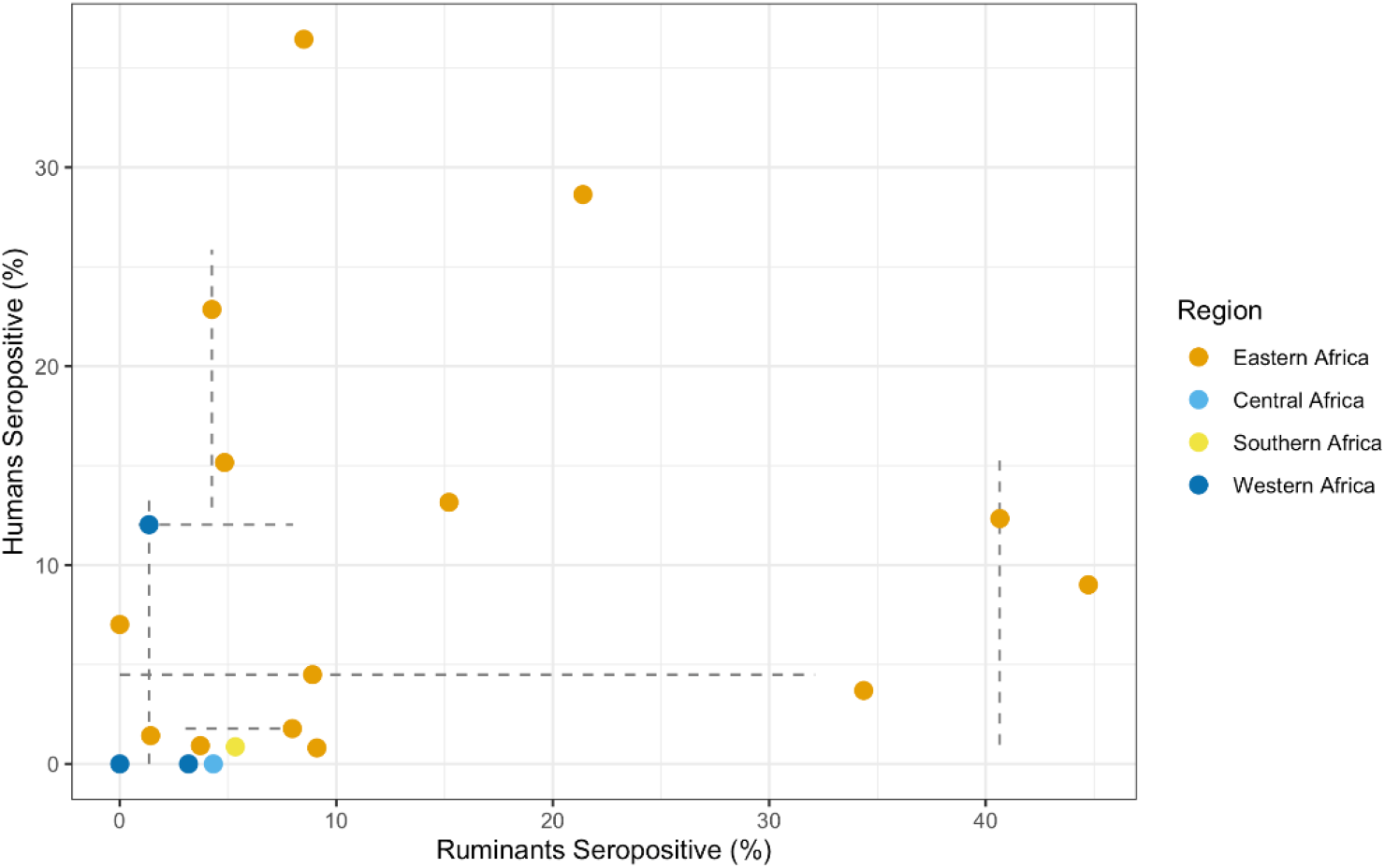
Animal and human RVF seroprevalence data from the same year and country. Point estimates represent mean seropositivity (percent of individuals testing positive of the total number of individuals tested), and lines represent the range in seroprevalence estimates reported when multiple publications were included in the prevalence estimate.

### 3.4. Concurrent animal and human RVFV activity

#### 3.4.1. Association of human RVF outbreaks with animal RVF cases

There were 74 concurrent human and animal RVF outbreaks (i.e., human and animal cases reported in the same year and country); 77% of 96 human outbreaks and 33% of 224 animal outbreaks occurred concurrently. Concurrent outbreaks were reported in 21 counties, with most in South Africa (15), Mauritania (9) and Kenya (7).

For half of the concurrent outbreaks case counts were available for both humans and animals (37 of the 74 paired records). The human and animal case counts of concurrent outbreaks (37 paired records) were weakly positively correlated (Spearman S=5910, rho=0.299, P=.07, Suppl. Figure 2). The occurrence of human RVF cases increased the likelihood that case counts were available for animal RVF records: 58% (43 of 74) of concurrent outbreaks and 30% (45 of 150) of animal-only outbreaks had case counts (OR: 3.2, 95%CI: 1.74, 6.02, P<.001).

Of the 22 human RVF outbreaks without associated animal RVF cases, 16 occurred within two years of an RVF outbreak in animals, leaving 6 human RVF outbreaks without a link to reported animal RVF cases (6.3% of 96 human RVF outbreaks: 150 cases in Kenya 2014 - 2015, 15 cases in Central African Republic 2019, 1 case in Gambia 2018, an unknown number of cases in Namibia 1974, and Uganda 1968).

#### 3.4.2. Correlation between animal and human seroprevalence

A total of 17,269 people (selected using a random sampling strategy) and 11,777 ruminants were sampled in the same year and country. The 19 paired records of concurrent seroprevalence data, include 10 countries (Kenya n=9, Senegal n=2, Central African Republic, Ethiopia, Madagascar, Mauritania, Mayotte, Tanzania, Uganda and South Africa) and 16 years (earliest 1982, most recent 2017).

Countries with high seroprevalence in animals were not typically associated with high human seroprevalence during the same year (Spearman S=720, rho=0.368, P=.12, Figure 6). When considering the seroprevalence records that selected for high-risk individuals (e.g., herders, slaughterhouse personnel), the association with ruminant seroprevalence was even weaker (12 paired country-year records, Spearman S=120, rho=0, P=1). The absence of a significant correlation between human and ruminant seroprevalence may, among other factors, result from the fact that not all records originated from the same ADM1-level. ADM1-level information was available for human and ruminant seroprevalence records of 10 paired country-year records. Of those, 3 country-year records included the same ADM1 for animal and human serological data, a spatial scale which more precisely reflects RVFV outbreaks.

## 4. Discussion

Approximately 90 years of reporting, 70 years of surveillance efforts, and about 20 years of near real-time case report availability allowed us to map the pathogen’s spread, assess the relative importance of common ruminant species sampled, and explore pathogen spillover to humans. Explorations of this patchwork of different data sources demonstrates the increase in available data on this pathogen. Analyses further indicate that exposure in animals and humans is likely to be increasing, and that sheep may play an important role in transmission of the pathogen, particularly during outbreaks. The data set generated (Suppl. Table 1) provides the first open-access, global, human and animal, RVF case and RVFV seroprevalence data compilation, and allows researchers to further investigate the epidemiology of the virus.

RVFV spread significantly over the past decades and appears to (re)emerge more frequently. For example, the pathogen spread to Saudi Arabia and Yemen from Africa in 2000. In addition, there is serological evidence of viral exposure in animals in Iraq [60], Iran [61] and Turkey [62] in the absence of RVF cases. However, as RVFV confirmation and animal travel histories are incomplete, local RVFV circulation cannot be established based on these records alone. Furthermore, the pathogen recently re-emerged in areas where no cases had been confirmed for over 40-50 years [63]. The increase in outbreak reports is in part due to improved reporting systems that are globally accessible, but not exclusively. Greater mobility and trade could facilitate movement of seropositive animals. These movements in combination with increased densities of humans and animals, changing mosquito population dynamics, landscapes and weather patterns could all contribute to faster introductions and subsequent transmission. Indeed, animal and human exposure to RVFV appear to have increased over time. The approximate 30% increase in seropositivity in humans and 6% increase in animals every 10 years (e.g., 5% seroprevalence becomes 6.5% or 5.35%), suggests increased exposure to the virus. However, these changes in seropositivity could in part be due to improved, more sensitive, diagnostic tools, and changed sampling strategies, whereby cross-sectional studies screening for hemorrhagic fever viruses were replaced by RVFV-specific studies in areas with known or suspected virus activity (e.g., outbreak investigations, intervention evaluations). In addition, an increase in seroprevalence is to be expected in longer-lived species, like humans, even if the force of infection is relatively stable over time, due to the accumulation of exposed individuals who survived infection (i.e., seropositive individuals). Age-stratified sampling could help disentangle these historical transmission patterns [64], but few studies reported on the age of participants and fewer provided age-stratified data (but, for examples see [65–67]). In addition, structured, long-term seroconversion studies that include both humans and main animal hosts could help provide local evidence of the force of infection and how it changes over time.

Our analyses corroborated a prominent role for sheep during outbreaks, finding them to be the most likely species to seroconvert during outbreaks, three times more likely than during cryptic transmission cycles. In comparison, goats were approximately 1.5 times more likely to seroconvert. This aligns with the belief that sheep are the most important host species for RVFV amplification, owing to their high susceptibility and viral loads [2,68]. These characteristics, in addition to sheep experiencing the most severe pathology due to RVFV, could make them the prime vaccine target to minimize economic losses for farmers and to prevent RVF outbreaks. Understanding the species’ local contribution to RVFV maintenance and amplification is important for vaccination and surveillance strategies to prevent RVF outbreaks.

Despite several large RVF outbreaks in animals, human case counts remained relatively low, suggesting either limited or geographical variation in spillover, or infections going unrecognized, undiagnosed, or unreported. In people, the probability of asymptomatic infection is estimated to range from 90 to 98% [69,70]. In addition, health care access and diagnostics may be limited, and RVF symptoms overlap with those of other prevalent diseases [71], possibly obscuring case identification and reporting. Similarly to humans, animal RVF cases may also be overlooked thereby facilitating cryptic transmission of RVFV. The absence of animal cases when human RVF cases were observed suggests that livestock cases also go unreported, or human cases were strictly wildlife and mosquito mediated. In a herd, RVF is easy to overlook when few animals are affected, or few animals show signs of disease [72]. As suggested by the greater likelihood of having case counts for animals when human cases occur simultaneously, outbreak investigations among animals are often initiated after human cases have been detected. In addition, the presence of RVFV antibodies without animal or human RVF cases in a country could be explained by cryptic transmission, whereby cases may be mild enough or in small enough numbers to go unnoticed. Some of the presence of antibodies in the absence of cases could be explained by RVFV exposure at a different geographical location, i.e., in traded animals, and seroconversion due to vaccination. Overall, the evidence of cryptic transmission urges us to reconsider reliance on passive surveillance systems for the detection of RVFV, and other emerging pathogens.

Increased data availability, through historical reviews, studies sharing their (raw) data, and near-real time reporting of RVF cases, facilitated the creation of this comprehensive data set, thereby connecting the patchwork of RVFV data and improving epidemiological inference made in these publications. Our study highlights the value of detailed reviews at a national level that summarize historic local and national database records - many previously unavailable - with as much detail as possible (for example [41,43]). Furthermore, our data set illustrates the importance of centralized databases, as many of RVF case records were sourced from ProMED-archives and the OIE. Notably, ProMED combines grey-literature with official reporting of cases and outbreaks to international organizations, expanding its reach beyond traditional data sources. We thus purposely expanded our search strategy to go beyond peer-reviewed published (experimental) studies (an integrated approach, e.g., including theses, ProMED, OIE databases), used a broad case definition, and accepted all serological tests detecting IgG. By adopting this strategy, we were able to picture a more complete view of the history of RVF epidemiology. However, the robustness of data and conclusions from the analyses should also be considered in this context. The information gained from patterns in the data should be further explored and investigated in more detail when additional data becomes available.

Standardization in reporting would strengthen the data set and the inferences that can be made from the data in future analyses. For example, serological studies were inconsistent in reporting the importation and vaccination history of the animals, possibly inflating seroprevalence. Similarly, the migration and travel history of people was often not included, placing the location of RVFV antibody detection away from the true location of infection. In addition, although districts, regions or provinces of sampling were often reported, the data was not shared at these geographical levels in about a quarter of publications. Therefore, we summarized data by country and year to create a more robust data set. This aggregation sacrifices the finescale information and prevents capturing within-country variation. Since RVF outbreaks are often localized, and they can prompt outbreak investigations and seroprevalence studies beyond the affected area; our aggregation of the data set combined studies from areas with and without RVFV transmission, thereby possibly underestimating seroprevalence associated with outbreaks. Furthermore, we identified samples as outbreak-associated when samples were collected during the outbreak, or the year after the outbreak (reported RVF cases in humans and/or animals). This grouping potentially placed samples collected at outbreak locations two years or more after an outbreak in the non-outbreak group (e.g., sampling in Egypt 13 years post an outbreak [73]). Combined, this meant that our adjusted odds ratios likely underestimated the actual increase in human and animal seroprevalence due to RVF outbreaks. As the geographical range suitable for RVFV introduction and establishment continues to expand, global efforts should continue to improve surveillance strategies to detect pathogen emergence and prevent pathogen spread. Similarly, with the increasing frequency of outbreaks, both in eastern and western Africa, regional and national programs may adjust their disease surveillance and RVFV intervention program. To evaluate if RVFV activity and exposure is increasing, as suggested by the increased seroprevalence over time, targeted long term studies in endemic and non-endemic areas could be started. These efforts would be most beneficial when a one health approach were to be used [74]: in which human, animal, and mosquito populations, and the environment are monitored simultaneously. Looking ahead, standardized reporting of serological studies, and uniform case definitions would ensure that on-the-ground, local efforts can be utilized at a larger geographical scale, further informing mathematical models and facilitating a thorough understanding of RVFV epidemiology. Ultimately, these research and reporting efforts combine local interests and international research priorities to limit the burden of RVFV.

## Data Availability

Data will be made available in Open Science Framework upon publication.

## Supplementary Materials

The following are available online Figure S1: Data availability over time (logistic regression plotted), Figure S2: Human and Animal RVF cases, Table S1: Data set, Table S2: Seroprevalence model output – People, Table S3: Seroprevalence mode output – Ruminant, Table S4: Seroprevalence model output - All animals, Table S5: Seroprevalence model output - Selected ruminant species

## Author Contributions

Conceptualization, GMB, QtB, QT, TAP; Methodology, GMB, KS, QtB, QT, TAP; Validation, HC; Formal Analysis, GMB; Investigation, GMB, KS; Data Curation, AL, GMB, KS; Writing – Original Draft Preparation, GMB, KS, QtB; Writing – Review & Editing, All authors; Visualization, GMB, KS, TAP; Supervision, QtB, SM, TAP; Funding Acquisition, GMB, QtB, TAP..

## Data Availability Statement

Data is contained within the supplementary material.

## Acknowledgments

We thank Robert Sumaye for his insightful comments and constructive review of the manuscript. We also thank several authors who kindly provided additional information regarding their studies upon request.

